# Roles of generation-interval distributions in shaping relative epidemic strength, speed, and control of new SARS-CoV-2 variants

**DOI:** 10.1101/2021.05.03.21256545

**Authors:** Sang Woo Park, Benjamin M. Bolker, Sebastian Funk, C. Jessica E. Metcalf, Joshua S. Weitz, Bryan T. Grenfell, Jonathan Dushoff

## Abstract

Inferring the relative strength (i.e., the ratio of reproduction numbers, ℛ_var_/ℛ_wt_) and relative speed (i.e., the difference between growth rates, *r*_var_ −*r*_wt_) of new SARS-CoV-2 variants compared to their wild types is critical to predicting and controlling the course of the current pandemic. Multiple studies have estimated the relative strength of new variants from the observed relative speed, but they typically neglect the possibility that the new variants have different generation intervals (i.e., time between infection and transmission), which determines the relationship between relative strength and speed. Notably, the increasingly predominant B.1.1.7 variant may have a longer infectious period (and therefore, a longer generation interval) than prior dominant lineages. Here, we explore how differences in generation intervals between a new variant and the wild type affect the relationship between relative strength and speed. We use simulations to show how neglecting these differences can lead to biases in estimates of relative strength in practice and to illustrate how such biases can be assessed. Finally, we discuss implications for control: if new variants have longer generation intervals then speed-like interventions such as contact tracing become more effective, whereas strength-like interventions such as social distancing become less effective.

## 1 Introduction

Estimating variant epidemic strength and speed remains one of the key questions in controlling the spread and understanding the threat of SARS-CoV-2 variants of concern (VoCs) (Althaus et al., 2021; Davies et al., 2021; Di Domenico et al., 2021; Graham et al., 2021; Leung et al., 2021; Volz et al., 2021; Zhao et al., 2021). Epidemic “strength” is measured by the reproduction number ℛ– the average number of new infections caused by a typical infection. Disease can spread in a population if ℛ*>* 1 (Diekmann et al., 1990). The epidemic strength also determines the final size of an epidemic in a homogeneous population (Anderson and May, 1991). Epidemic “speed” is characterized by the growth rate *r*, which describes how fast a disease spreads at the population level. Like epidemic strength, epidemic speed also determines conditions for disease elimination: *r* = 0 is a threshold equivalent to ℛ= 1. Strength and speed are linked by generation intervals—defined as the time between infection and transmission—which characterizes the individual-level time scale of the epidemic (Roberts and Heesterbeek, 2007; Svensson, 2007; Wallinga and Lipsitch, 2007).

Analyses of new variants have typically characterized *relative* strength (i.e., the ratio of reproduction numbers ℛ_var_/ℛ_wt_) and speed (i.e., the difference between growth rates *r*_var_ −*r*_wt_) of the variants. Many studies have tried to estimate the relative strength of the novel variant from the observed relative speed (Davies et al., 2021; Leung et al., 2021; Volz et al., 2021; Zhao et al., 2021). Some studies have instead assumed a value of the relative strength of the novel variant and tried to predict its relative speed to determine when a new variant will become the dominant strain (Davies et al., 2021; Di Domenico et al., 2021). While both approaches are reasonable, holding different quantities constant (i.e., strength or speed) can lead to different conclusions about the spread of the disease and its control (Dushoff and Park, 2021): estimation of speed from strength or strength from speed depends on assumptions about the generation-interval distribution (Roberts and Heesterbeek, 2007; Svensson, 2007; Wallinga and Lipsitch, 2007). For example, when epidemic strength is fixed, assuming longer generation intervals lead to a slower epidemic (lower *r*), making the epidemic look easier to control with speed-like interventions such as contact tracing. When epidemic speed is fixed, assuming longer generation intervals lead to a stronger epidemic (higher ℛ), making the epidemic look harder to control with strength-like interventions such as social distancing.

In practice, most studies have assumed that the previous dominant strain (wild type) and variants have identical generation intervals, but recent evidence suggests that the B.1.1.7 variant may have a longer duration of infection: 13.3 days (90% CI: 10.1–16.5) for the new variant and 8.2 days (90% CI: 6.5–9.7) for the wild type (Kissler et al., 2021). Longer duration of infection suggests that the mean generation interval of B.1.1.7 is likely to be longer than that of the wild type. Other studies have considered the possibility that the faster growth rate of new variants may be driven, in part, by shorter generation intervals (Davies et al., 2021; Volz et al., 2021). In general, if the generation-interval distribution of a new variant is different from the generation-interval distribution of the wild type, estimates of variant strength may be biased. However, linking strength and speed is complicated given that generation intervals depend on many factors including behavior: for example, self-isolation after symptom onset will lead to shorter generation intervals.

Here, we explore how different assumptions affect estimates of the relative strength ℛand speed *r* for a new variant, such as B.1.1.7. We compare the relationship between relative strength (the ratio ℛ_var_/ℛ_wt_) and relative speed (the difference *r*_var_ −*r*_wt_) under a wide range of assumptions about generation-interval distributions. We find that neglecting differences in the generation-interval distributions can lead to biased estimates. We also discuss how such biases might be assessed in practice and how information on differences in generation interval distributions might influence priorities for controlling the spread of VoCs.

## 2 Renewal equation framework

We use the renewal equation framework to characterize the spread of two pathogen strains— in this case, the wild type SARS-CoV-2 virus and a focal variant of concern. In particular, we focus on characterizing the incidence of infection (i.e., the rate at which new infections are generated), which is directly related to *r* and ℛ. In contrast, observed case reports are subject to reporting delays, which must be taken into account in order to correctly infer *r* or ℛ in practice (Goldstein et al., 2009; Gostic et al., 2020).

Neglecting the (relatively slow) rate of new mutations, the current incidence of infection *i*_*x*_(*t*) caused by each strain *x*—either the wild type (“wt”) or the variant (“var”)—can be expressed in terms of their previous incidence *i*_*x*_(*t − τ*) and the rate at which secondary infections are generated at time *t* by individuals infected *τ* time units ago *K*_*x*_(*t, τ*):

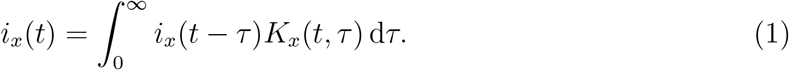

This framework provides a flexible way of modeling disease dynamics and generalizes compartmental model, such as the SEIR model (Heesterbeek and Dietz, 1996; Diekmann and Heesterbeek, 2000; Roberts, 2004; Aldis and Roberts, 2005; Roberts and Heesterbeek, 2007; Champredon et al., 2018).

The integral of the kernel ℛ_*x*_(*t*) = ∫*K*_*x*_(*t, τ*) d*τ* is referred to as the instantaneous reproduction number (Fraser, 2007). The instantaneous reproduction number is a particular kind of weighted average of infectiousness of previously infected individuals at time *t*—in particular, it is weighted by the total relative infectiousness at time *t*, rather than by the actual number of infected individuals present. The normalized kernel *g*_*x*_(*t, τ*) = *K*_*x*_(*t, τ*)/ℛ_*x*_(*t*)— which we refer to as the instantaneous generation-interval distribution—describes the relative contribution of previously infected individuals to current incidence *i*_*x*_(*t*) and provides information about the time scale of disease transmission. Both the reproduction number and the generation-interval distribution depend on many factors, including intrinsic infectiousness of an infected individual, non-pharmaceutical interventions, awareness-driven behavior, and population-level susceptibility (Fraser, 2007).

Over a short period of time, we can assume that epidemiological conditions remain roughly constant: ℛ_*x*_(*t*) ≈ ℛ_*x*_ and *g*_*x*_(*t, τ*) ≈ *g*_*x*_(*τ*). In this case, the incidence of each strain will exhibit exponential growth (or decay) at rate *r*_*x*_, satisfying the Euler-Lotka equation (Wallinga and Lipsitch, 2007):

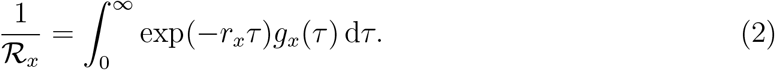

We can approximate this *r*– ℛ relationship by assuming that the generation-interval distribution is Gamma-distributed, and summarizing it using the mean generation interval 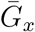 and the squared coefficient of variation *κ*_*x*_ (Park et al., 2019):

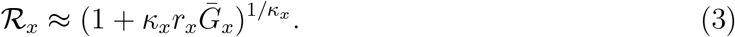

The Gamma assumption includes as a special case models that assume exponentially distributed generation intervals (when *κ* = 1), corresponding to the SIR model (Anderson and May, 1991); various Gamma assumptions are widely used in epidemic modeling, including for models of SARS-CoV-2 (Park et al., 2020a). We use this framework to investigate how inferences about strength and speed of the variant depend on our assumptions about the underlying generation-interval distributions. For simplicity we neglect differences in the squared coefficient of variation and assume *κ*_wt_ = *κ*_var_ = *κ*; instead, we focus on the effect of potential differences in the mean generation intervals.

## 3 Inferring relative strength from relative speed

Epidemic speed *r*_*x*_ can often be estimated directly from incidence of infection during the exponential growth period (Mills et al., 2004; Nishiura et al., 2009; Ma et al., 2014). However, estimating *r*_*x*_ can be challenging when case counts are low (as when a new variant begins to spread) and is sensitive to temporal changes in the distribution of testing effort in the population and the overall testing intensity. Studies of new SARS-CoV-2 variants have mostly focused on characterizing changes in the *proportion* of a new variant (Althaus et al., 2021; Davies et al., 2021; Di Domenico et al., 2021; Graham et al., 2021; Leung et al., 2021; Volz et al., 2021; Zhao et al., 2021). Focusing on proportions has the advantage, because changes in proportions are less sensitive to changes in testing and to other transient effects that would affect variants and wild type viruses similarly. When incidence is changing exponentially (*i*_*x*_(*t*) = *i*_*x*_(*t*_0_) exp(*r*_*x*_*t*)), the proportion of the new variant *p*(*t*) follows a logistic growth curve:

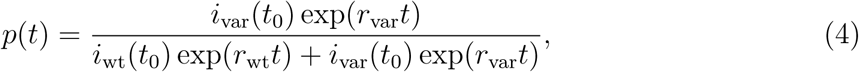

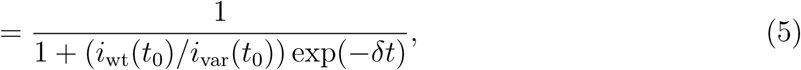

where the logistic growth rate *δ* = *r*_var_ *−r*_wt_ corresponds to the relative speed of the epidemic.

We thus ask: what factors affect the relative strength *ρ* = ℛ_var_/ℛ _wt_ of a new variant, conditional on an observed relative speed *δ*? Inference of *ρ* from *δ* will depend on assumptions about the generation-interval distributions of both strains. Given the mean generation interval of the variant 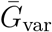 and the wild type 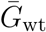, the relative strength *ρ* = ℛ_var_/ℛ _wt_ under the Gamma assumption (Park et al., 2019) is given by:

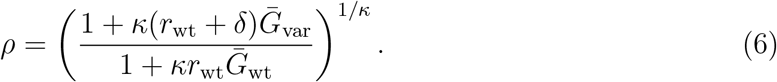

Therefore, the relative strength *ρ* depends not only on the relative speed *δ* and the generation-interval distributions but also on how fast the wild type is spreading in the population (*r*_wt_)— some analyses have implicitly or explicitly neglected this factor by either assuming *r*_wt_ = 0 (Althaus et al., 2021) or *κ* = 0 (Davies et al., 2021) (in the latter case 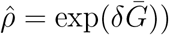.

Based on the observed relative growth rate of B.1.1.7 in the UK, we start by taking the relative speed of the variant to be *δ* = 0.1*/*day (Davies et al., 2021); the mean generation interval of the wild type to be 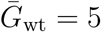days (Ferretti et al., 2020b); and the squared coefficient of variation of generation intervals to be *κ* = 0.2 (Ferretti et al., 2020b). We consider the spread of B.1.1.7 as an example but our qualitative conclusions should hold for other VoCs.

As noted above, we assume that the variant and the wild type have equal *κ* throughout, and only consider differences in the mean. We evaluate the estimates of relative strength *ρ* across a wide range of *κ* from 0 (fixed-length generation intervals) to 1 (exponential distribution). To further explore how inference depends on underlying epidemiological conditions (i.e., whether the incidences of the variant and the wild type are growing or shrinking), we consider five epidemiological conditions: (1) *r*_wt_ *< r*_var_ *<* 0, (2) *r*_wt_ *< r*_var_ = 0, (3) *r*_wt_ *<* 0 *< r*_var_, (4) 0 = *r*_wt_ *< r*_var_, and (5) 0 *< r*_wt_ *< r*_var_.

Fig. 1 illustrates how the relative strength depends on the shape of the generation-interval distributions and on underlying epidemiological conditions. In general, longer mean generation intervals of the new variant translate to higher values of *ρ* (and vice versa), except when *r*_var_ *≤* 0 (recall, we always assume *r*_wt_ *< r*_var_). When *r*_var_ = 0, we always have ℛ_var_ = 1 and so the inferred value of *ρ* is independent of the generation-interval distribution of the new variant. When *r*_var_ *<* 0, we see that longer generation intervals decrease *ρ* because longer generation intervals actually lead to slower decay (higher *r*). Assuming a narrower distribution (lower *κ*) has qualitatively similar effects as assuming longer generation intervals because both reduce the amount of early transmission; therefore, narrower distributions lead to higher values of *ρ* when *r*_var_ *>* 0 and lower values of *ρ* when *r*_var_ *≤* 0. When *r*_wt_ *<* 0 *< r*_var_, inference of *ρ* is relatively insensitive to values of *κ*.

**Figure 1:**
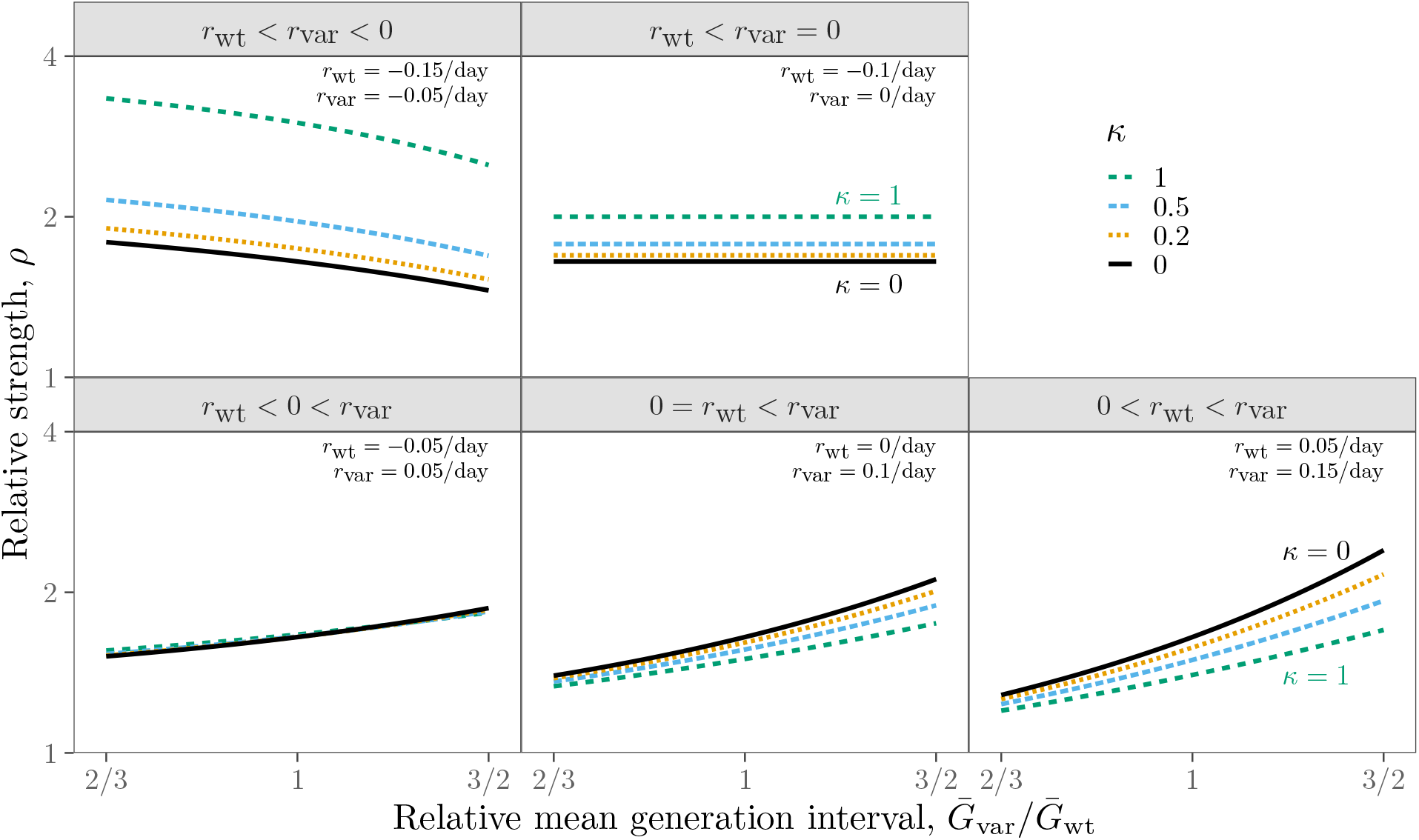
Relative strength of the new variant assuming a fixed speed advantage *δ* under five epidemiological conditions. The relative strength of the new variant *ρ* conditional on the speed of the wild type *r*_wt_; the ratio between the mean generation interval of the new variant 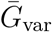 and that of the wild type 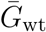; and the squared coefficient of variation in generation intervals *κ*. The relative strength of the new variant *ρ* is calculated using *δ* = 0.1*/*day, 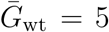days, and *κ* = 1*/*5. Assumed values of *r*_wt_ (and therefore *r*_var_) are shown in the top right corners of each panel.

Unsurprisingly, we find that an increased speed of *δ* = 0.1*/*day for the variant is consistent with higher strength than the wild type across the range of epidemiological conditions considered (Althaus et al., 2021; Davies et al., 2021; Di Domenico et al., 2021; **?**; Graham et al., 2021; Leung et al., 2021; Volz et al., 2021; Zhao et al., 2021). However, the magnitude of relative strength *ρ* is sensitive to assumptions about generation intervals. For realistic values of *κ* (excluding 0 and 1), we find that the inferred relative strength *ρ* ranges between 1.1–2.3. Even if we restrict the difference in generation interval to less than 20% and only consider scenarios with *r*_var_ *>* 0 (bottom panels of Fig. 1), we find that *ρ* ranges between 1.4–1.8.

## 4 Inferring relative speed from relative strength

We do not generally expect the relative speed *δ* to remain constant if other factors governing epidemic spread are changing. Instead, many biological mechanisms appear compatible with assuming a constant value of relative strength *ρ* over changing conditions. For example, if the proportion of the population susceptible declines, or the average contact rate changes, while other factors remain constant, the relative strength *ρ* is expected to remain constant; in general, this will imply a change in relative speed *δ*.

We thus investigate how *δ* is expected to change with ℛ _wt_ if *ρ* remains constant, and how this expectation changes with the ratio of the generation intervals. Once again, we rely on the Gamma assumption to find the relative speed *δ* given the mean generation interval of the variant 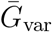 and the wild type 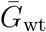,:

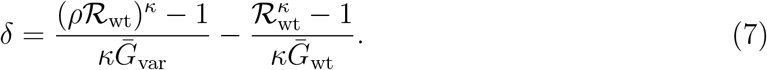

As our baseline scenario we assume *ρ* = 1.61, which is the value we obtain for *δ* = 0.1*/*day, *r*_wt_ = 0*/*day, 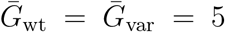days, and *κ* = 1*/*5. We evaluate *δ* across five different epidemiological conditions as before: (1) ℛ_wt_ < ℛ_var_ *<* 1, (2) ℛ_wt_ < ℛ_var_ = 1, (3) ℛ_wt_ *<* 1 < ℛ_var_, (4) 1 = ℛ_wt_ < ℛ_var_, and (5) 1 < ℛ_wt_ < ℛ_var_.

In general, longer generation intervals lead to slower relative speed of the variant when the incidence of both strains is increasing (Fig. 2, bottom panels) because slower growth of the variant reduces the differences in absolute speed. When ℛ_var_ = 1, the relative speed is insensitive to the generation-interval distribution of the variant because we always have *r*_var_ = 0. When ℛ_var_ *<* 1, longer generation intervals of the variant lead to slower decay (*r*_var_ closer to 0), and therefore, greater relative speed. Once again, we see that assuming a narrower distribution (lower *κ*) has qualitatively similar effects as assuming a longer mean.

**Figure 2:**
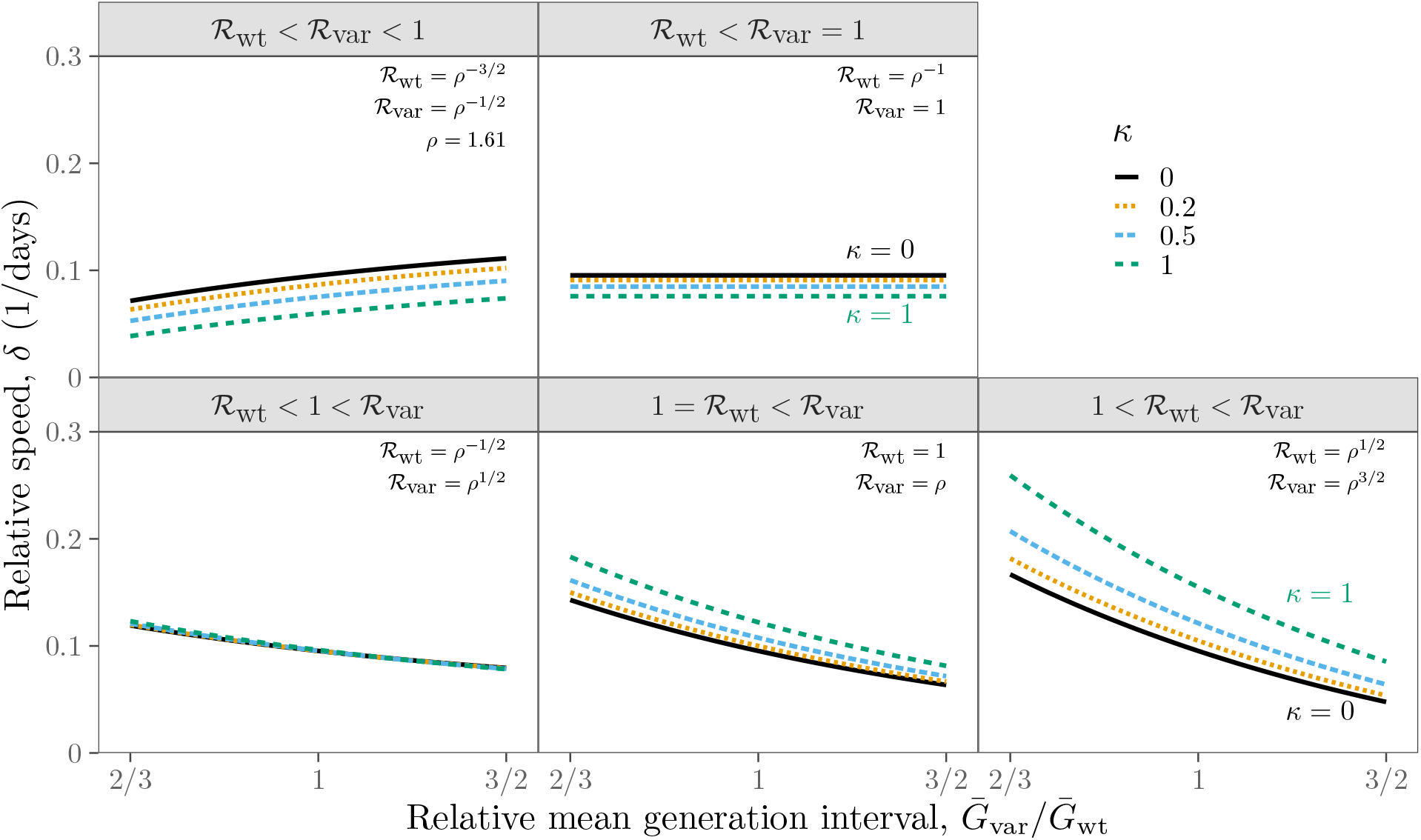
Relative speed of the new variant assuming a fixed strength advantage *ρ* under five epidemiological conditions. The relative speed of the new variant *δ* conditional on the strength of the wild type ℛ_wt_; ratio between the mean generation interval of the new variant 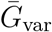 and that of the wild type 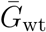; and squared coefficient of variation in generation intervals *κ*. Relative speed of the new variant *δ* is calculated using *ρ* = 1.61, 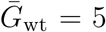days, and *κ* = 1*/*5. Assumed values of ℛ_wt_ (and therefore ℛ_var_) are shown in the top right corners of each panel.

**Figure 3:**
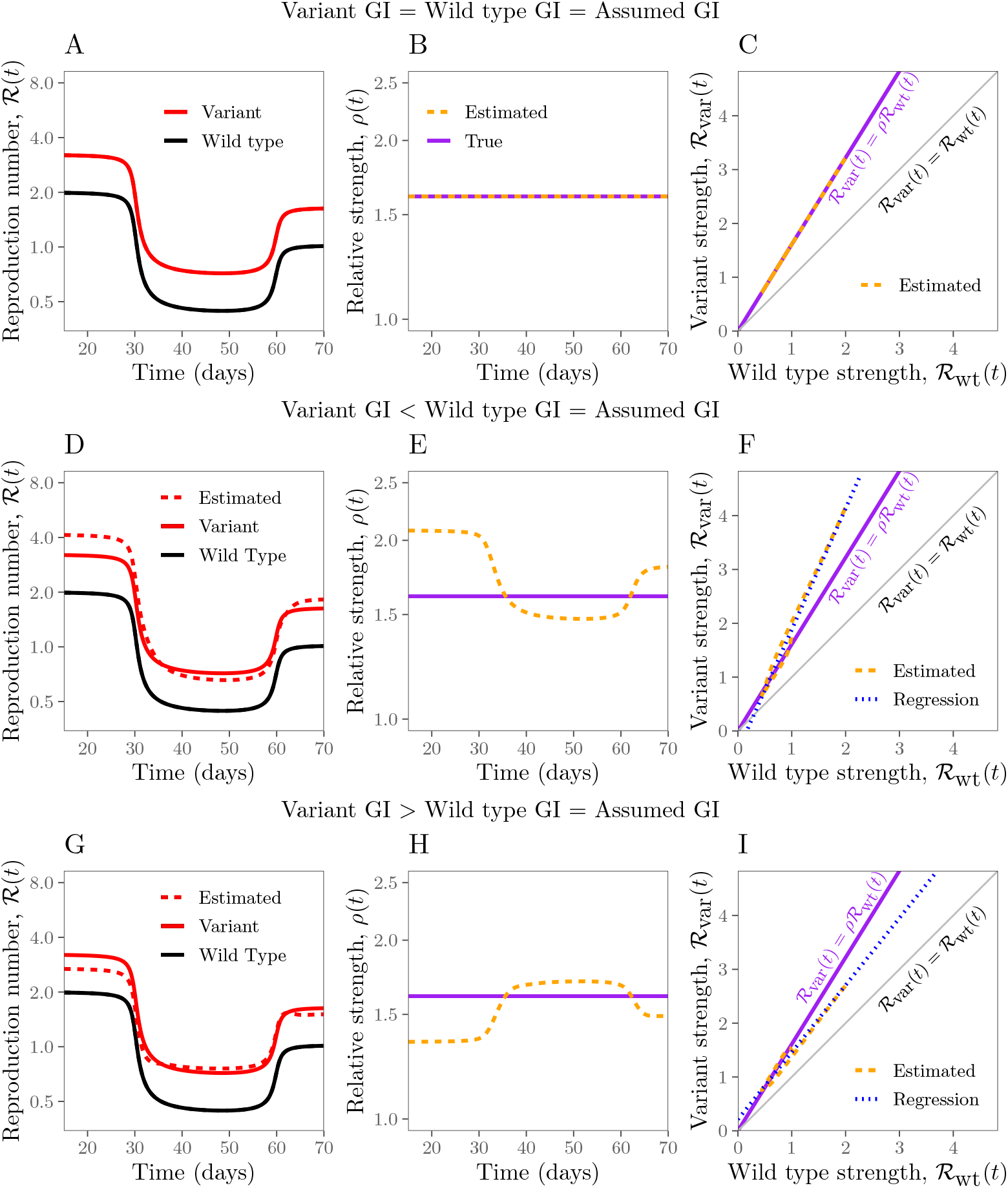
Estimates of relative strength over time under different scenarios. (A, D, G) True (solid lines) and estimated (dashed lines) reproduction numbers of the new variant and the wild type over time. (B, E, H) True (purple, solid) and estimated (orange, dashed) ratios between reproduction numbers of the new variant and the wild type over time. (C, F, I) Phase planes (time is implicit) showing true (purple, solid) and estimated (orange, dashed) relationships between estimated reproduction numbers. Blue dotted lines represent the regression lines of the estimated variant reproduction numbers against the estimated wild type reproduction numbers. Gray lines represent the one-to-one line. Top row: the assumed mean generation-interval (5 days) is equal to the mean generation-interval of the wild type and the variant. Middle row: the assumed mean generation-interval (5 days) is equal to the mean generation-interval of the wild type but is longer than that of the variant (4 days). Bottom row: the assumed mean generation-interval (5 days) is equal to the mean generation-interval of the wild type but is longer than that of the variant (6 days). For all simulations, squared coefficient of variation in generation intervals is assumed to equal *κ* = 1*/*5.

Fig. 2 also shows that when *ρ* is fixed relative speed depends on underlying epidemiological conditions. For example, even when there are no differences in the generation-interval distributions (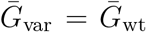, in this case), the relative speed *δ* can range between 0.08–0.11 when *κ* = 0.2 and 0.06–0.14 when *κ* = 0.5. Therefore, characterizing the spread of variants assuming constant relative speed (e.g., by fitting a standard logistic growth curve) without considering how epidemiological conditions change over time should be used with care.

## 5 Inferring relative strength from incidence data

Instead of estimating relative strength from speed, one can estimate time-varying or instantaneous reproduction numbers ℛ(*t*) of the variant and the wild type from incidence data (Fraser, 2007), and directly compare their ratios; such methods have been used in previous analyses of the B.1.1.7 strain by Volz et al. (2021). Assuming that the generation-interval distribution remains constant, the instantaneous reproduction numbers of the new variant and of the wild type can be estimated from their corresponding incidence curves—an approach popularized by Cori et al. (2013):

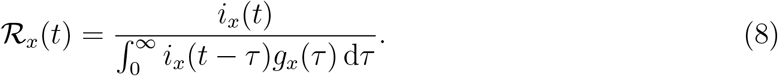

Under strength-like intervention measures that reduce transmission rates by a constant amount, we expect ratios between reproduction numbers to remain constant and correspond to the true relative strength: ℛ _var_(*t*)/ℛ_wt_(*t*) = *ρ*. However, if the assumed generation-interval distribution *ĝ*(*τ*) is different from the true distribution, then the ratio between the estimated reproduction numbers 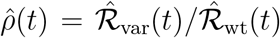 may change, even if the true ratio does not.

Here, we investigate how misspecification of the generation-interval distribution of the variant affects our inference of relative strength from inference data under the assumption that the true generation-interval distribution of the wild type is known. We use a two-strain renewal equation that assumes perfect cross-immunity to simulate three different scenarios (see Methods): (1) the wild type and the variant have the same generation-interval distributions (which match the known distribution, shown in Fig. 5A–C); (2) the variant has a shorter mean generation interval (Fig. 5D–F); and (3) the variant has a longer mean generation interval (Fig. 5G–I). Then, we compare the estimated ratio 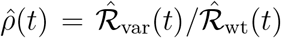 with the true ratio *ρ* = ℛ_var_(*t*)/ℛ_wt_(*t*). In order to allow for changes in ℛ driven by the introduction and lifting of non-pharmaceutical interventions, we let ℛ_wt_(*t*) decrease from 2 to 0.4 around day 30 and increase back up to 1 around day 60 and assume ℛ_var_(*t*) = *ρ ℛ*_wt_(*t*). Although previous studies have modeled the impact of non-pharmaceutical interventions as a step function (Flaxman et al., 2020), we choose to use a smooth function to model ℛ_wt_(*t*) (Fig. 5; see Methods) given the possibility that behavioral changes may affect transmission before and after interventions take place. We reach similar conclusions if we use a step function instead (Supplementary Figure S1).

When the assumed distribution matches the true distribution, the estimated reproduction numbers match the true values (Fig. 5A); thus, their ratio remains constant and 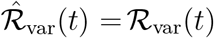 (Fig. 5B). However, if the generation-interval distribution of the variant differs from the assumed distribution, the ratio changes over time (Fig. 5E,H). In particular, if the true generation intervals of the variant have a shorter mean than the assumed distribution, we over-estimate ℛ _var_(*t*) during the growth phase and under-estimate during the decay phase (and conversely, Fig. 5D,G), which further translates to biases in the estimated relative strength (Fig. 5E,H).

In practice, estimates of instantaneous reproduction numbers ℛ (*t*) (and therefore, their ratios) can be noisy due to limited data availability or model assumptions; instead, we might want to estimate a single value of relative strength *ρ*. For example, we can estimate *ρ* by plotting the estimated strength of the variant 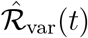 against the estimated strength of the wild type 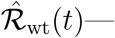—as presented in Figure 2 of Volz et al. (2021)—and performing a linear regression (Fig. 5C,F,I). If *ρ* is constant, and generation-interval distributions are correctly specified, we obtain a straight line with a slope of *ρ* and intercept at zero (Fig. 5C). However, when the assumed mean generation interval is longer than the that of the variant, we over-estimate the slope (and conversely, Fig. 5F,I). Biases in slopes further translate to biases in intercepts; in theory, we would expect the regression line to go through the origin (because ℛ_var_ = 0 when ℛ_wt_ = 0).

## 6 Implications for intervention strategies

While relative speed *δ* and strength *ρ* are useful for characterizing the spread of the variant in an epidemiological context with a previously dominant wild type, the *absolute* speed *r*_var_ and strength ℛ _var_ of the variant are the main variables that determine the spread and conditions for control of the variant over the long term. In particular, at any given point in the epidemic, we can measure the speed of the variant *r*_var_ (or infer *r*_var_ from *r*_wt_ and *δ*) and ask how much more intervention is required to control the spread of both strains (since ℛ_var_ *<* 1 implies ℛ_wt_ *<* 1). As a baseline scenario, we assume *r*_wt_ = 0 and *δ* = 0.1*/*day (and therefore *r*_var_ = 0.1*/*day), in which case additional intervention is required to reduce *r*_var_ below 0 (or, equivalently, ℛ_var_ below 1).

We consider two types of intervention: an intervention of constant strength, which reduces transmission by a constant factor *θ* regardless of age of infection (*K*_post_(*τ*) = *K*_pre_(*τ*)*/θ*); and an intervention of constant speed, which reduces transmission after infection by a constant rate *ϕ* (*K*_post_(*τ*) = *K*_pre_(*τ*) exp(*−ϕτ*)). In this case, we can control the spread of the variant when *θ > ℛ*_var_ or *ϕ > r*_var_, respectively (Dushoff and Park, 2021). We consider constant-strength and speed interventions that reduce ℛ_var_ to 0.9 when both the variant and the wild type have equal mean generation intervals (Fig. 4A–B). While both interventions are equally effective on the strength scale (that is, ℛ_post_ = ∫*K*_post_(*τ*) d*τ* = 0.9), they have different dynamical implications. The constant-strength intervention affects transmission equally throughout the course of infection, whereas the constant-speed intervention has greater impact on transmission that occurs later in infection; as a result, the constant-speed intervention reduces the post-intervention mean generation interval (Fig. 4B) and leads to (slightly) faster exponential decay (therefore, lower *r*_post_).

**Figure 4:**
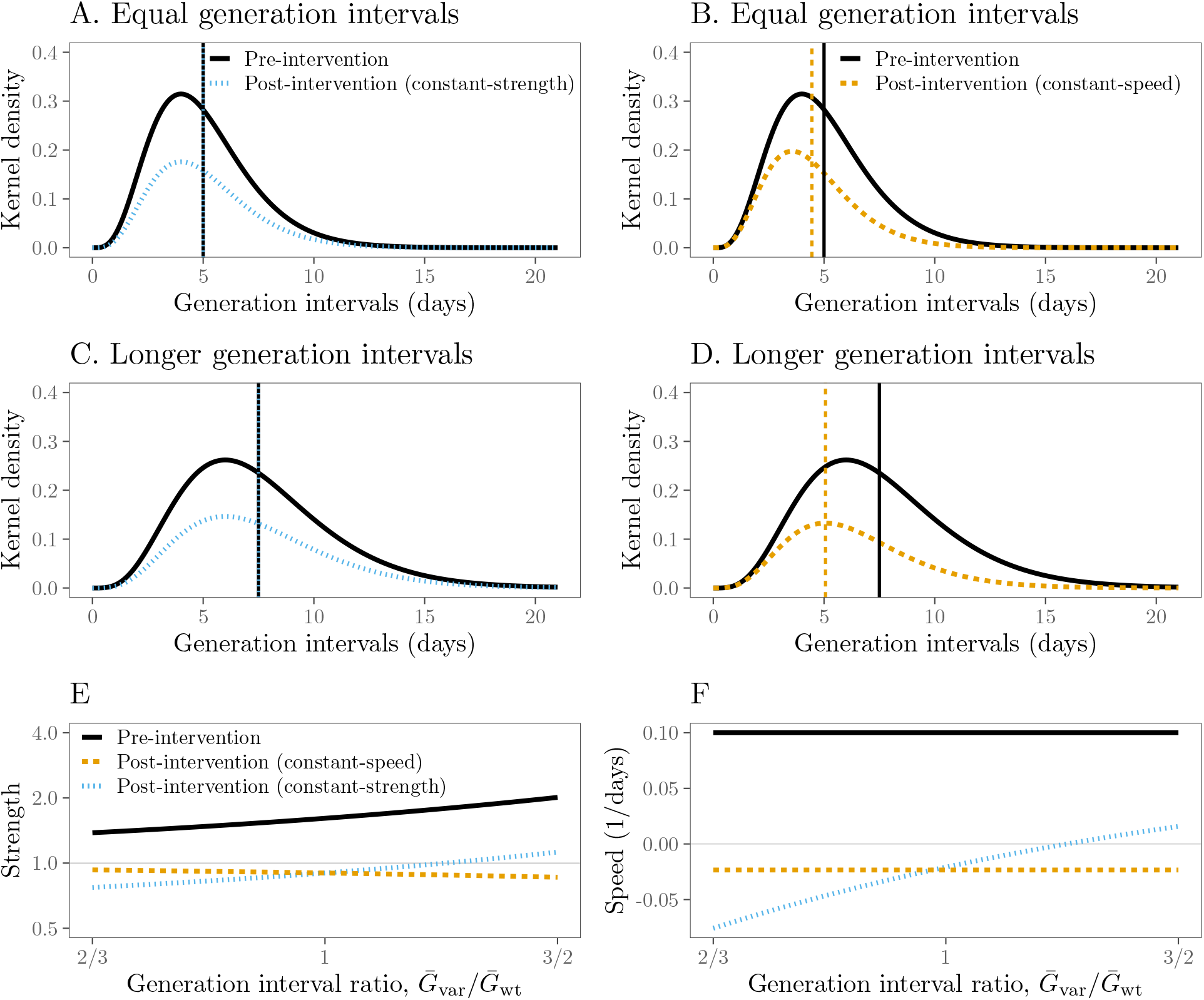
Effects of constant-strength and constant-speed interventions on the spread of a new variant with known speed *r*_var_. (A–B) Pre-intervention (black) and post-intervention (colored) kernel of the new variant under constant-strength (A) and –speed (B) interventions assuming that the variant has equal generation intervals as the wild type. (C–D) Pre-intervention (black) and post-intervention kernel of the new variant assuming that the variant has longer generation intervals than the wild type. (E) Pre-intervention (black) and post-intervention strength conditional on the mean generation interval of the variant (colored). (F) Pre-intervention (black) and post-intervention speed conditional on the mean generation interval of the variant (colored). Strength and speed are calculated assuming *r*_wt_ = 0*/*day, *δ* = 0.1*/*day, *r*_var_ = 0.1*/*day, 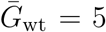days, and *κ* = 1*/*5 for pre-intervention conditions. Intervention strength and speed are chosen so that post intervention strength of the new variant is 0.9 when its mean generation interval is 5 days.

However, if the variant has longer generation intervals than the wild type (Fig. 4C–D), then the strength of the variant will be higher conditional on the observed speed (Fig. 4E).

In this case, the same constant-strength intervention can fail to control the epidemic (i.e., ℛ_post_ *>* 1; Fig. 4E) because this intervention reduces the transmission by a constant amount regardless of age of infection (Fig. 4C). On the other hand, the same constant-speed intervention will prevent a larger proportion of transmission, leading to lower ℛ_post_ (Fig. 4E), because it is more effective against late-stage transmission (Fig. 4D). The constant-speed intervention also reduces the mean generation interval by a larger factor (Fig. 4D).

The speed-based paradigm gives the same results regarding control but provides important insights (Fig. 4F). The observed speed of the variant *r*_var_ at a given moment is independent of our estimates of its mean generation interval. Likewise, the post-intervention speed of the variant under the constant-speed intervention is also independent of the mean generation interval; therefore, the constant-speed intervention will always be equally effective (on the speed scale) in preventing the spread.

The constant post-intervention speed can be understood by considering a speed-based decomposition of the kernel: *K*_pre_(*τ*) = exp(*r*_var_)*b*(*τ*), where *b*(*τ*) is the initial backward generation-interval distribution and therefore integrates to 1 (Dushoff and Park, 2021). This decomposition allows us to re-write the post-intervention kernel (under the constant-speed intervention) as follows:

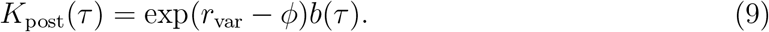

Since the post-intervention speed *r*_post_ is determined by the following implicit equation,

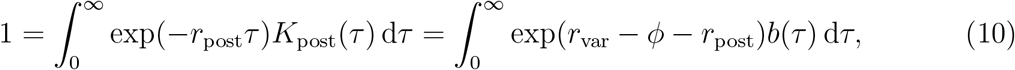

we have *r*_post_ = *ϕ − r*_var_. In particular, if speed of intervention is faster than the observed speed of spread (i.e., if *ϕ > r*_var_), we can control the epidemic (i.e., *r*_post_ *<* 0) regardless of the underlying generation-interval distribution.

## 7 Discussion

We explored how the generation-interval distribution shapes the link between relative strength and speed of an invading disease variant. Longer generation intervals generally lead to higher relative strength for a given relative speed (and conversely, lower relative speed for a given relative strength); these relationships are reversed when incidence is decreasing. Neglecting potential differences in the mean generation intervals between the variant and the wild type can bias estimates of the relative strength from incidence data; these biases may be assessed by considering whether relative strength appears to vary systematically with underlying epidemiological dynamics. Finally, differences in generation intervals can also lead to different conclusions about the effectiveness of interventions: if the variant has longer generation intervals than the wild type, speed-like interventions, will be relatively more effective than naive estimates would suggest (and conversely, strength-like intervention will be relatively more effective if the variant has shorter generation intervals).

As new variants, in particular the B.1.1.7 variant, of SARS-CoV-2 are spreading and becoming dominant in many countries, it is clear that the variants are more transmissible (have higher strength) than the wild type (Althaus et al., 2021; Davies et al., 2021; Di Domenico et al., 2021; Graham et al., 2021; Leung et al., 2021; Volz et al., 2021; Zhao et al., 2021). Our analysis supports estimates of a higher strength of the new variant across a wide range of assumptions about epidemiological conditions and generation-interval distributions. However, our analysis also shows that uncertainty in generation-interval distributions must be taken into account to obtain accurate estimates of the relative strength of variants. If new variants has a longer infectious period (Kissler et al., 2021), and therefore extended generation-interval distributions, current estimates may underestimate the true relative strength. If new variants escape vaccinal or natural immunity to some extent (Kupferschmidt, 2021), their relative strength may increase further.

There is currently a (minor) discrepancy in the estimates of relative strength of new variants, particularly for B.1.1.7. Mathematical analyses have typically reported greater than a 1.4 fold increase in reproduction number for the B.1.1.7 variant (*ρ >* 1.4) whereas an independent analysis of secondary attack rates from contact tracing data suggests a 1.25–1.4 fold increase (Public Health England, 2020). As shown in Davies et al. (2021) and Volz et al. (2021), propagating uncertainty in generation interval estimates (and, in particular, assuming shorter generation intervals) can partially explain these differences.

Using the Gamma-approximation framework (Park et al., 2020a) can help us reconcile different estimates more clearly. For example, Davies et al. (2021) estimated a relative strength of *ρ* = 1.77 assuming a fixed generation generation interval at 5.5 days from *δ* = 0.104*/*day in one of their regression analysis. Similarly, Volz et al. (2021) estimated *ρ* = 1.79 by directly comparing the ratio between ℛ_var_ and ℛ_wt_ assuming a mean generation interval of 6.5 days and a squared coefficient of variation of 0.38. In this case, the latter study’s longer mean generation interval (which increases *ρ*) is counter-balanced by the effects of their wider distribution (which decreases *ρ*); calculations using our approximation framework produce qualitatively similar results (*ρ* = 1.81 from 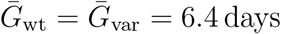, *κ* = 0.38, *δ* = 0.104*/*day, and *r*_wt_ = 0*/*day). If we consider short and wide generation interval estimates from Tianjin, China (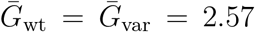 and *κ* = 1; Ganyani et al. (2020)), we obtain *ρ* = 1.27 (from *δ* = 0.104*/*day and *r*_wt_ = 0*/*day). Although these estimates are more consistent with the attack rate analysis (Public Health England, 2020), we do not claim that our estimates of *ρ* are more accurate. This calculation simply highlights the importance of propagating uncertainty in generation-interval distributions in assessing the relative strength and speed of SARS-CoV-2 variants.

We further used simulations to show how mis-specification of generation-interval distributions can bias the inference of relative strength from incidence data: when the assumed generation-interval distribution does not match the true generation-interval distribution of the variant, estimates of absolute strength ℛ _var_(*t*) will be biased, causing the estimated relative strength to change over time. In our simulations, we assumed that the intervention would reduce transmission caused by the variant and the wild type by equal amounts, thereby preserving the relative strength over time; however, this assumption only holds under strength-like interventions, which are insensitive to age of infection, but not under speed-like interventions. As we demonstrated, the effectiveness of speed-like interventions depends on the shape of the generation-interval distribution; therefore, if the variant and the wild type have different generation intervals, speed-like interventions such as contact tracing can affect them differently.

We also assumed that the incidence of infection caused by the variant and the wild type is known. However, estimating ℛ (*t*) from real data, such as case incidence, is often associated with a variety of practical challenges, including delays between infection and reporting and changes in testing patterns (Gostic et al., 2020). We have chosen to focus here on the underlying dynamical mechanisms that may affect inference. Future studies may consider the development of methods based on mechanism outlined here to assess potential biases in their estimates of relative strength.

In moving from mechanism to improved estimates, we argue that both perspectives (i.e., whether we hold relative strength or speed constant) are useful in understanding the dynamics of the new variant. Early in the spread of an emerging variant, it is likely more convenient to fix the relative speed, given that speed can be directly observed. As more information about the transmission and immunity profiles of the new variant becomes available, we advise instead fixing the relative strength and inferring the speed, as this assumption better matches biological mechanisms for the variants’ higher strength (e.g., higher rates of transmission and immune evasion). Researchers should be mindful about which quantity they hold constant and how their conclusions follow from their assumptions (Dushoff and Park, 2021).

This study has practical implications for analyzing the epidemiological dynamics of new variants. First, models that assume a constant relative speed, such as the standard logistic growth model, should be used with care—it is important to remember that the relative speed is expected to change with epidemiological conditions. In some cases, such as early in the spread of new variants, assuming a constant relative speed may be justified if epidemiological conditions remain relatively constant. Such simplifying assumption can also help obtain crude estimates of relative speed. However, epidemiological conditions *will* change over time in response to spreading new variants, causing relative speed to change and invalidating this assumption. Second, the absolute strength and speed should not be neglected in favor of relative values; relative and absolute values should be given roughly equal weight when presenting conclusions. While the relative strength and speed are useful for describing the spread of new variants, the absolute values determine their spread and control. Finally, uncertainty in generation intervals should be carefully considered.

Even though SARS-CoV-2 has been spreading for more than a year, there is still considerable uncertainty about its generation intervals. Observational studies typically focus on serial intervals (i.e., time between symptom onset of the infector and the infectee; Svensson (2007)) because they are easier to measure (Griffin et al., 2020). However, realized serial intervals are subject to dynamical biases that can be difficult to tease apart (Park et al., 2021). A few studies have tried to estimate the generation-interval distribution from serial intervals, with means ranging between 3–6 days and squared coefficients of variation ranging between 0.1–1 (Ferretti et al., 2020b,a; Ganyani et al., 2020; Knight and Mishra, 2020); as we have shown, this uncertainty in the generation-interval distribution alone can generate large uncertainty in the inference of relative strength. Furthermore, most estimates neglect dynamical biases in serial intervals caused by underlying epidemiological dynamics in their estimates of generation intervals (Park et al., 2021). Serial intervals also fail to account for asymptomatic transmission, adding further uncertainty to inferences of speed and strength (Park et al., 2020b).

Since the beginning of the SARS-CoV-2 pandemic, a few studies have emphasized the relevance of leveraging generation interval distributions to improve estimates of strength (Park et al., 2020a; Ali et al., 2020; Gostic et al., 2020; Park et al., 2021) and the relative importance of modes of transmission (e.g., asymptomatic vs. symptomatic, Park et al. (2020b)). These same lessons should be considered when assessing the spread of variants in a partially susceptible population and improving efforts to control spread. Future studies should prioritize detailed assessment of the generation intervals of SARS-CoV-2 and widespread variants, as well as consider how uncertainty in generational intervals might bias conclusions.

The spread of new SARS-CoV-2 variants and the replacement of previously dominant lineages represent ongoing challenges for controlling the SARS-CoV-2 pandemic (Abdool Karim and de Oliveira, 2021; Fontanet et al., 2021; Walensky et al., 2021). By explicitly considering epidemiological context and generation interval differences together, we have shown that improving estimates of the the relative duration of infectiousness at the individual scale may represent a pathway towards more effective interventions. Specifically, speed-like interventions, such as contact tracing, will be relatively more effective if variants have longer generation intervals. Most intervention strategies throughout the current pandemic have focused on strength-like interventions (Flaxman et al., 2020), such as lock-downs, partly because pre-symptomatic transmission of SARS-CoV-2 has limited the effectiveness of contact tracing efforts (Hellewell et al., 2020). However, given the possibility that new variants can have different infection characteristics (Kissler et al., 2021), future studies should consider whether their transmission dynamics also differ (e.g., the amount of pre-symptomatic transmission) and evaluate intervention strategies accordingly.

## 8 Methods

### 8.1 Two-strain renewal equation

We use a two-strain renewal equation to simulate the spread of the variant and the wild type. Ignoring birth and death, the incidence of infection caused by the variant *i*_var_ and the wild type *i*_wt_ assuming perfect cross immunity is given by:

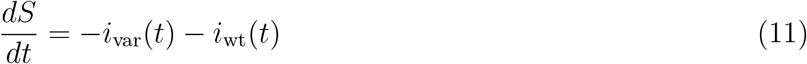

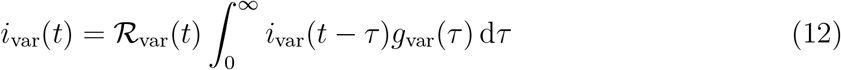

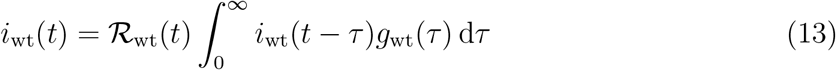

where *S* represents the proportion of susceptible individuals. We discretize the model at the time scale of 0.025 days as described in Park et al. (2021) and simulated the model with *i*_var_(0) = 0.001 and *i*_wt_(0) = 0.1. To allow for smooth changes in ℛ_wt_(*t*) around day 30 and 60, we let:

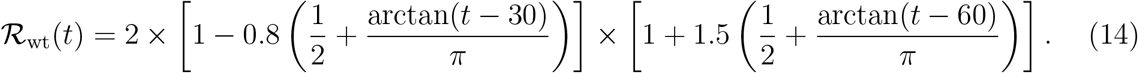

In this case, ℛ_wt_(*t*) remains around 2 and slowly decreases to 0.4 = 2 *×* (1 *−* 0.8) around day 30, and slowly increases back up to 1 = 0.4 *×* (1 + 1.5) around day 60. Finally, we assume ℛ_var_(*t*) = ρ ℛ_wt_(*t*). From simulated incidence curves between day 15 and 70, we estimate the instantaneous reproduction number using Eq. 8. We ignore incidence before day 15 to remove any potential transient effects.

## Data Availability

All data and code are stored in a publicly available GitHub repository (https://github.com/parksw3/newvariant)

## Supplementary Figure

**Figure S1:**
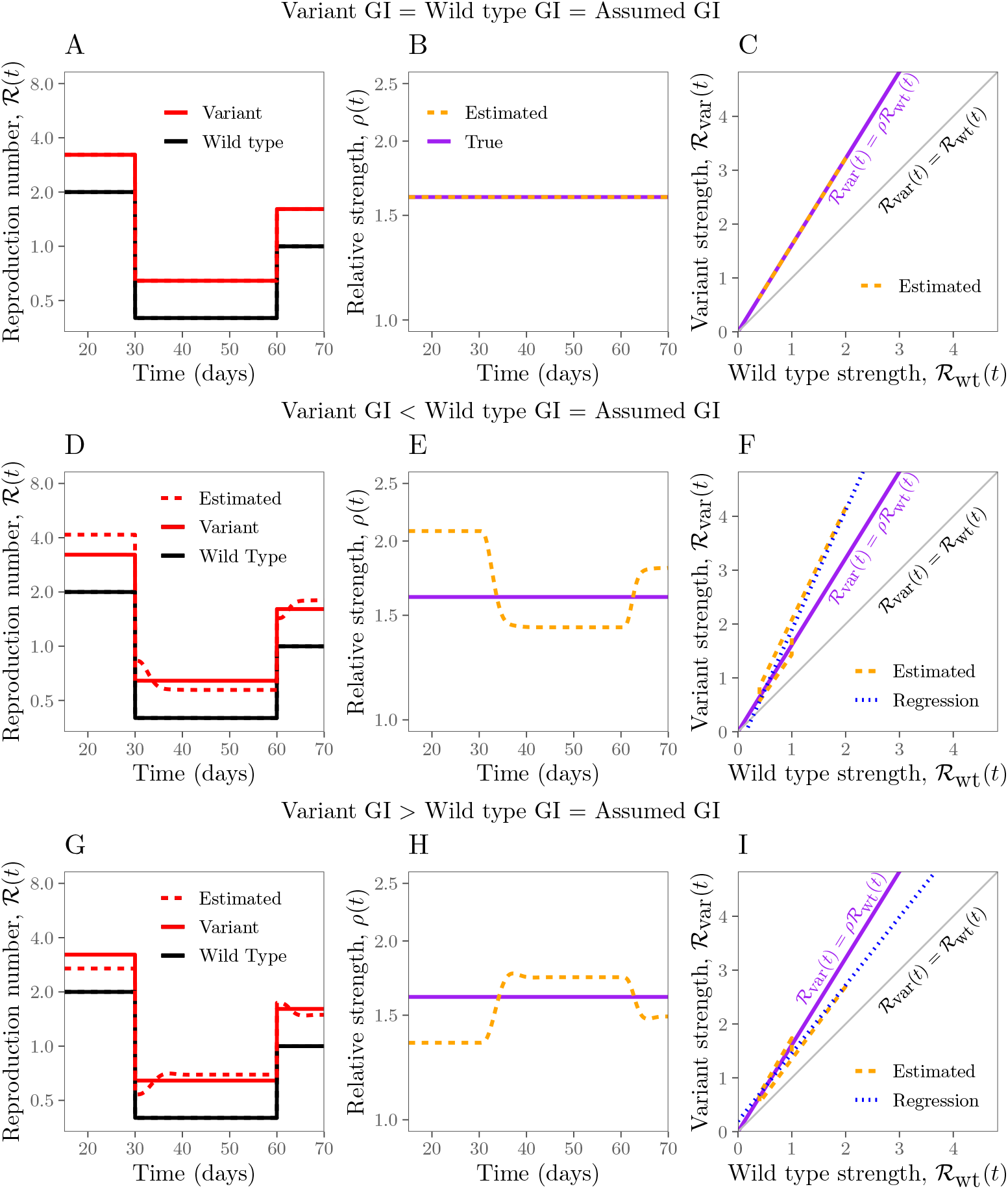
Estimates of relative strength over time under different scenarios assuming step changes in reproduction number. See Figure 3 of the original text for figure caption.

